# Uncovering the Molecular Landscape of Physical Activity: Proteomic Insights from the UK Biobank

**DOI:** 10.1101/2025.06.27.25330418

**Authors:** Hanyu Qian, Chao Wu, Yupeng Yang, Bo Li, Anthony Rosenzweig, Meng Wang

**Author notes:** Joint first author (Hanyu Qian, Chao Wu).

## Abstract

Different patterns of physical activity (PA), including sedentary behavior, have been linked to a wide range of health outcomes. However, the underlying molecular mechanisms— particularly the biological pathways connecting distinct aspects of PA to health outcomes—are incompletely understood. In this study, we investigated the associations between 14 PA features across four categories—captured using wearable accelerometer data—and plasma protein (PP) expression levels in 9,210 individuals from the UK Biobank. Our analysis identified 2,655 significantly associated PA feature–PP expression pairs, involving 613 unique proteins. These included broadly associated proteins such as ITGAV and MYOM3, which were linked to all PA feature categories, as well as proteins uniquely associated with specific activity features. Mediation analysis revealed 359 paths linking 10 PA features, 99 unique PPs, and seven incident health outcomes. Aggressive PA features, including both the proportion of moderate-to-vigorous physical activity (MVPA) time and the frequency of MVPA bouts, exhibited protective effects mediated through proteins to those risks for incident health outcomes. Proteins such as GDF15, PRSS8, and IGFBP4 mediated associations across multiple PA features. Additionally, genetic analyses identified 32 mediator proteins with putative causal effects on incident health outcomes, highlighting them as potential therapeutic targets. Together, these results implicate PP as key mediators of the health effects of PA, offering new mechanistic insights and uncovering potential targets for therapeutic intervention.

## Introduction

Physical activity (PA), including sedentary behavior, has been linked to a wide range of health outcomes. Insufficient PA and prolonged sedentary behavior are well-established risk factors for multiple chronic conditions, including cardiovascular disease, diabetes, and neurodegenerative disorders^1,2^. With the growing availability of wearable sensor technologies, it is currently feasible to derive a rich set of objective PA features, which enhances measurement accuracy, reduces bias, and facilitates more nuanced investigation into the relationship between PA and health outcomes^3,4^. While the duration of moderate-to-vigorous physical activity (MVPA) remains a primary focus in epidemiological studies, recent evidence suggests that additional aspects of PA behavior, such as the frequency and duration of sedentary bouts and intensity of short activity breaks, also contribute meaningfully to cardiometabolic health^5–8^. These studies highlight that multidimensional PA features may provide more biological insights than MVPA alone and therefore hold potential for revealing uncharacterized biological pathways linking behavior to diseases.

Circulating plasma proteins (PPs) offer a promising molecular avenue for exploring the relationship between PA and health outcomes^9,10^. A prior study using data from the Framingham Heart Study identified five PPs associated with PA with a limitation that the targeted panel of 1,305 proteins mainly focused on cardiovascular pathways^11^. More recent efforts leveraging large-scale proteomic data from the UK Biobank (UKB) have expanded this scope, profiling over 2,900 plasma proteins in 53,026 individuals. However, these studies primarily rely on subjective PA data from self-reported questionnaires ^12^ or are restricted to limited accelerometer-derived features, such as MVPA and average acceleration^13^. While these pioneering studies have advanced our understanding of the relationship between PA and health outcomes, it remains unclear whether associations between PPs and PA vary across specific behavioral dimensions of PA, and whether such differences correspond to distinct disease-relevant molecular mechanisms. A multidimensional set of PA features derived from high-resolution wearable accelerometer data may offer deeper insights into the PP links between PA and health outcomes.

In this study, we built a set of 14 objectively measured PA features derived from wrist-worn accelerometer data collected over a week in a free-living environment from 9,210 UK Biobank participants and further analyzed their associations with PP profiles^14,15^. These 14 PA features spanned four categories (average intensity, proportional activity status, state transition probability, and bout characteristics), offering a multidimensional representation of habitual PA behaviors. In this study, we leveraged circulating PPs to explore how diverse dimensions of PA relate to health outcomes (Fig. 1). We identified 613 plasma proteins significantly associated with at least one PA feature, including 237 proteins linked to features from all four categories, highlighting substantial heterogeneity in protein associations across PA dimensions. Mediation analyses revealed that 99 proteins may mediate the relationship between PA and incident health outcomes. Further, Mendelian randomization supported potential causal effects in 32/99 of these proteins. By characterizing the molecular signatures associated with distinct aspects of PA, our findings provide insights into the biological mechanisms through which PA influences health and highlight candidate biomarkers for future prevention and intervention strategies.

**Figure 1.**
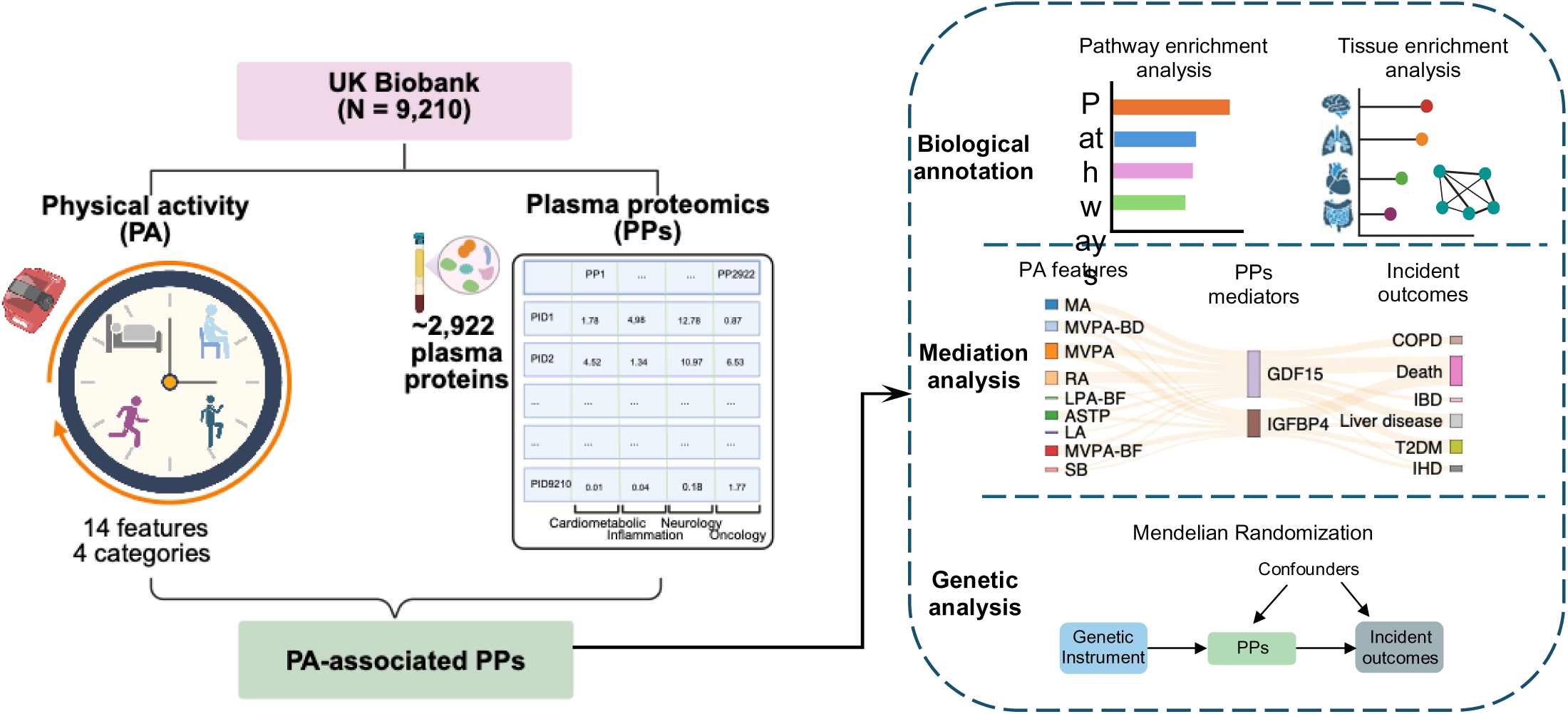
Schematic overview of the study design. The association analysis was conducted using data from 9,210 UK Biobank participants who completed both accelerometer-based physical activity (PA) assessment and plasma proteomic profiling. Fourteen PA features were derived and categorized into four dimensions to capture a comprehensive representation of PA patterns. Proteins associated with these PA features were further analyzed to annotate their biological functions, evaluate their potential mediating roles in the associations between PA and incident health outcomes, and investigate their potential causal effects on diseases through genetic analysis. Figure created with BioRender.com

## Result

### PPs and PA Population Characteristics

The discovery cohort included participants from UKB who enrolled in both the wrist-worn accelerometer substudy^16^ and the Olink Explore 3072 platform proteomics substudy ^10^. After data preprocessing (Methods), we retained 9,210 individuals with complete PA features, well-quantified expression levels for 2,922 PPs, and health outcome records for downstream analysis. The mean age of participants was 56.8 years (standard deviation SD = 8.0), and 56% were female. Detailed participant characteristics are provided in Supplementary Table 1.

### Comprehensive PA Features Derived from Accelerometer Data

PA features were derived from minute-resolution accelerometer data ^16^. To capture distinct dimensions of PA, we defined 14 PA features summarized into four categories: (i) Average intensity, including the mean log acceleration during the least active 6-hour window (LA; 12:00–6:00 AM) and the most active 18-hour period (MA; 6:00 AM–12:00 AM), along with relative amplitude (RA = (MA − LA) / (MA + LA)) (Fig. 2a) ^17^; (ii) Proportional activity status, reflecting the average proportion of daily time spent in sedentary behavior (SB; <30 milli-g), light physical activity (LPA; 30–100 milli-g), and moderate-to-vigorous physical activity (MVPA; >100 milli-g) ^14,18^; (iii) Transition probabilities (TP), quantifying the likelihood of transitioning from sedentary to active states (SATP) and from active states to sedentary (ASTP), where “active” encompassed both LPA and MVPA ^17^; and (iv) Bout characteristics, including mean duration and frequency of uninterrupted SB, LPA, and MVPA episodes, representing sustained activity patterns ^18^. The 6:00 AM threshold was selected based on empirical data, where activity levels remained consistently low before 6:00 AM and sharply increased thereafter (Supplementary Fig. 1, details in Method section).

**Figure 2.**
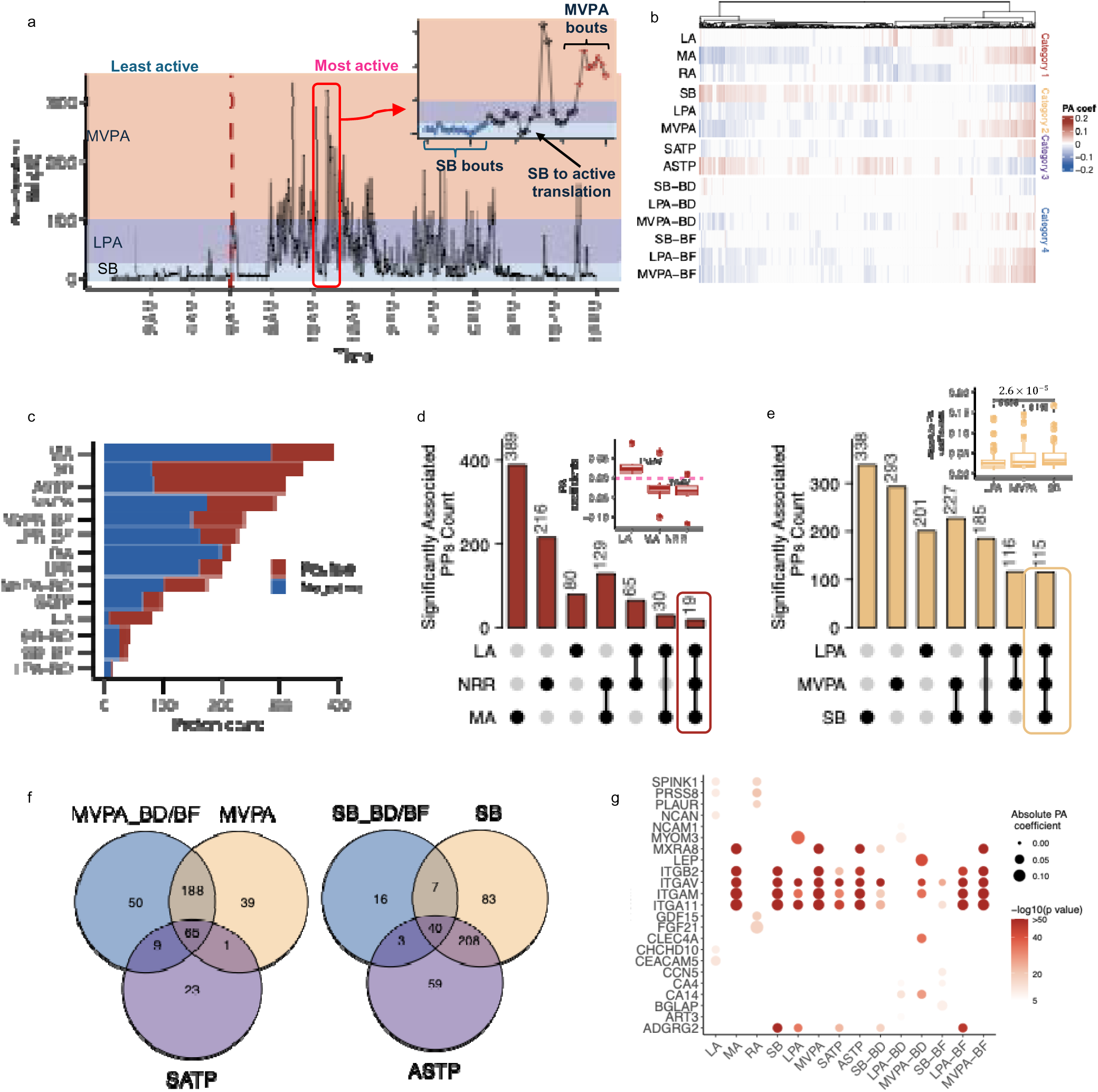
Associations between physical activity (PA) features and plasma protein (PP) expression (a) Example of acceleration data from one participant, with an illustration of the four PA feature categories. The four categories were average intensity, proportional activity status, transition probabilities, and bout characteristics (b) Heatmap depicting the coefficients of PA features with PPs expression, where non-significant coefficients are penalized to zero. (c) Number of PPs significantly associated with each PA feature, with positive associations in red and negative associations in blue. (d) UpSet plot of PPs associated with LA, MA and RA, and boxplot of linear model coefficients for PPs commonly associated with LA, MA, and RA. (e) UpSet plot of PPs associated with SB, LPA and MVPA and boxplot displaying the coefficients of the proportion of LPA MVPA and SB time in relation PPs expressions. The pairwise comparisons are from the t-test. (f) Venn diagrams showing the overlap of PPs associated with different categories of PA features.. (g) Dot plot illustrating the top five PPs most strongly associated with each PA feature.

Average intensity features captured overall activity levels during the LA and MA periods of the day, enabling a clearer delineation of rest–activity alternations than 24-hour averages. This segmentation is biologically relevant, as elevated activity during LA may indicate rest disruption, with potential implications for PP expression and health outcomes. RA, defined as the normalized contrast between MA and LA, reflected rest–activity alternations with lower values suggesting irregular activity patterns. While proportional activity status (i.e. time spent in SB, LPA, and MVPA) captured overall activity distribution, it did not account for behavioral dynamics. Therefore, we included TP features, quantifying SATP and ASTP, to reflect behavioral fragmentation and inertia. Bout characteristics, including the mean duration and frequency of continuous SB, LPA, and MVPA episodes, further described the temporal structure of PA. Together, TP and bout metrics provided complementary insights into the complexity and stability of daily activity patterns in relation to PP expression and disease risk.

### PA Features Show Varied Associations with PPs

We assessed associations between 14 PA features spanning four categories and the expression levels of 2,922 PPs using linear models adjusted for age, sex, and body mass index (BMI). A total of 2,655 PA–PP associations were significant under Bonferroni correction (p < 1.71 × 10□□= 0.05/2,922) ^19^. Overall, 613 PPs were associated with at least one PA feature (Fig. 2b; Supplementary Tables 2 and 3).

We observed widespread associations between PA features and PP expression levels, with some proteins linked to multiple PA features across categories. Average intensity features (LA, MA, and RA) were associated with the largest number of unique PPs (n = 480), suggesting strong molecular signatures of overall activity patterns. This was followed by proportional activity status (n = 419), bout characteristics (n = 383), and transition probabilities (n = 343). Among individual features, the MA metric—capturing average log acceleration over the most active 18-hour period—was linked to the greatest number of proteins (n = 389), of which 26.2% were positively and 73.8% negatively associated (Fig. 2c). Other top informative features included SB proportion (338 PPs) and ASTP (310 PPs). Within bout metrics, MVPA-BF exhibited the strongest associations (n = 240), underscoring the biological relevance of frequent vigorous activity. Overall, sedentary features were more likely to show positive associations with protein expression, whereas high-intensity or dynamic features—including MA, RA, MVPA duration and frequency, SATP, and MVPA time—were predominantly linked to decreased protein levels (Fig. 2c).

Nineteen PPs were significantly associated with all three average intensity features (LA, MA, and RA) with 18 displaying inverse associations (positive with LA, negative with MA), indicating consistent molecular signatures of disrupted rest–activity balance (Fig. 2d). Integrin alpha-V (ITGAV) was the sole exception, showing positive associations with all three metrics (Fig. 2d). Of the 216 PPs associated with RA, 41 were not identified through LA or MA alone (Supplementary Table 2), suggesting RA captures unique sensitivity to relative activity contrast rather than absolute intensity. In the proportional activity domain, 115 PPs were jointly associated with SB, LPA, and MVPA, typically exhibiting positive associations with SB and negative associations with active behaviors (Fig. 2e). Paired *t*-tests on the absolute association coefficients (|β|) indicated that the associations of SB and MVPA with PP expression were significantly stronger than LPA. Although SB showed the largest average effect size, its difference from MVPA was not statistically significant, suggesting that both prolonged sedentary time and vigorous activity may elicit stronger molecular responses than light activity.

Bout features related to MVPA—specifically, bout duration (MVPA-BD) and frequency (MVPA-BF)—were significantly associated with 312 PPs, including 59 not identified through MVPA time alone (Fig. 2f). Notable examples among these newly identified proteins include Carboxypeptidase A2 (CPA2), a metabolic enzyme associated with MVPA-BF (r = 0.033, *p* = 1.57 × 10□□, r was the linear model coefficient), and Hepatitis A virus cellular receptor 1 (HAVCR1), linked to MVPA-BD (r = −0.040, *p* = 4.82 × 10□□). The ASTP, which quantifies the likelihood of reverting to sedentary behavior, revealed 62 additional PPs beyond those associated with sedentary time alone. Among these, NTRK3 showed the strongest association (r = −0.016, *p* = 5.47 × 10□¹¹). These results highlight the additional benefits of incorporating dynamic PA metrics, such as bout characteristics and transition probabilities, for capturing nuanced biological signatures of distinct activity patterns.

A total of 237 PPs were significantly associated with at least one PA feature from each of the four feature categories. ITGAV exhibited the highest number of associations (n = 13, excluding LPA-BD), consistent with the established role of integrins in mechanotransduction, particularly within cardiovascular and musculoskeletal systems^20^. To highlight proteins with the strongest associations, we selected the top five PPs per PA feature (Fig. 2g), revealing involvement in pathways central to PA and exercise adaptation. These included regulators of muscle structure and repair (e.g., MYOM3, integrins) ^21^, metabolic homeostasis (e.g., LEP, FGF21, GDF15, osteocalcin)^22–24^, immune-mediated tissue remodeling (e.g., ITGB2, ITGAM, CLEC4A) ^25–27^, and acid–base balance (e.g., CA4, CA14)^28^. MYOM3, a sarcomeric protein essential for muscle integrity, showed strong associations across multiple features (e.g., SB: *r* = – 0.168, *p* = 1.67 × 10□□□; MVPA: *r* = 0.127, *p* = 1.38 × 10□³□; ASTP: *r* = –0.123, *p* = 2.01 × 10□³□). Leptin (LEP), a key metabolic hormone, was associated with eight PA features and exhibited the largest effect size with MVPA time, consistent with its role in energy regulation and exercise-induced leptin sensitivity. Additional proteins implicated in ECM remodeling, immune modulation (e.g., PLAUR, MXRA8, CCN5, CLEC4A), and mitochondrial or acid–base regulation (e.g., CHCHD10, CA4, CA14) may reflect molecular pathways underpinning exercise-driven physiological adaptations.

### Biological function of the PA-associated proteins

To understand the biological functions of identified PPs associated with each PA feature, we performed pathway enrichment analysis using the GO:BP ^29,30^ and KEGG ^31^ database to annotate the associated biological processes and pathways ^32^.

Our analysis revealed that cytokine-related pathways, the PI3K-Akt signaling pathway, and phagosome pathways were significantly enriched among the proteins associated with various PA features (11 out of 14) (Fig. 3a-d, and Supplementary Fig. 2, Table 4 and 5). This suggests that regular activity, including SB, may modulate key biological processes such as immune cell activation, inflammatory signaling, cellular survival, and tissue remodeling. The PI3K-Akt pathway, in particular, is well known for its role in regulating muscle hypertrophy, glucose homeostasis, and cell growth, all of which are critically influenced by levels of PA^33^. Similarly, phagosome-related pathways indicate potential links to immune surveillance and cellular debris clearance, processes that could be elevated in response to exercise-induced muscle microdamage and subsequent repair.

**Figure 3.**
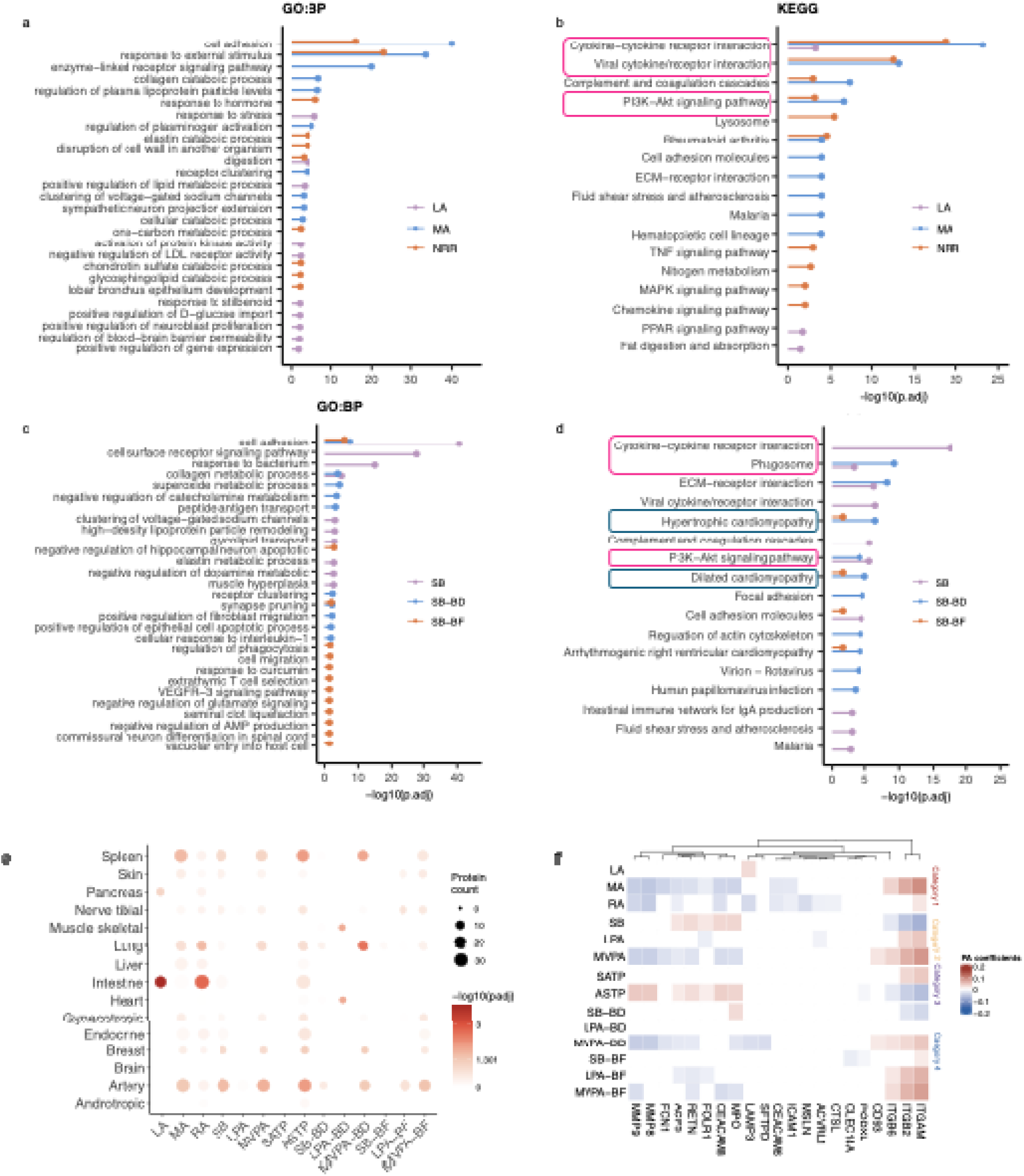
Biological function annotation of physical activity (PA)-associated plasma proteins (PPs). (a –b) Pathway enrichment analysis results for PPs associated with LA, MA, and RA using Gene Ontology Biological Processes (GO:BP) and KEGG pathways, respectively. (c - d) Pathway enrichment analysis results for PPs associated with SB, SB-BD, and SB-BF using GO:BP and KEGG pathways, respectively.(e) Number of enriched PPs in each tissue system and p values from the proportion test. (f) Heatmap showing the linear model coefficients between PA features and lung-enriched PP expression.

Notably, SB-BD associated proteins showed significant enrichment in pathways implicated in hypertrophic cardiomyopathy, dilated cardiomyopathy, and arrhythmogenic right ventricular cardiomyopathy (Fig. 3d and Supplementary Table 4 and Table 5). These findings align with clinical observations indicating that a sedentary lifestyle, characterized by prolonged sitting and minimal physical exertion, is a notable risk factor for the development of cardiovascular disease (CVD), including heart failure and increased mortality risk^34,35^. Taken together, these enrichment patterns highlight the critical role of immune regulation, cellular growth pathways, and cardiac remodeling processes in mediating the physiological and pathophysiological outcomes of varying PA levels.

### Tissue enrichment of the PA-associated PPs

We investigated whether the PA-associated PPs are tissue-enriched. We grouped the tissue-enriched proteins across 32 human tissues in the GTEx project into 15 functional systems (Methods and Supplementary Table 6) ^36–38^. In total, 242 PA-associated PPs were enriched in at least one tissue system. Enrichment proportions for each PA feature were compared to the full proteomic set (n = 2,922) using Fisher’s exact test. Significant enrichment (Benjamini– Hochberg-adjusted *p* < 0.05) was observed in the lung, intestine, spleen, and artery—tissues with well-established physiological links to physical activity and known for harboring functionally specialized proteins ^39–43^ (Fig. 3e). Notably, MMP8, MMP9, and FCN1—proteins implicated in lung development and fibrosis ^44,45^ —were associated with MA, RA, ASTP, and MVPA-BD (Fig. 3f), suggesting a possible role of PA in modulating pulmonary molecular pathways. Although not statistically enriched, 45 and 39 PA-associated PPs were also linked to the endocrine system and brain, respectively.

PPs associated with different categories of PA features exhibited distinct tissue enrichment patterns (Supplementary Fig. 3). For instance, proteins linked to the proportion of MVPA time were most enriched in the artery, whereas those associated with MVPA bout duration (MVPA-BD)—which extends MVPA proportion by incorporating temporal structure—were enriched in the lung (12 PPs) and spleen (16 PPs), reflecting divergent physiological signatures. These findings suggest that different PA feature types capture distinct dimensions of behavior, each linked to specific tissue systems. Thus, incorporating a comprehensive set of PA features is essential for capturing the full behavioral spectrum and elucidating the biological systems most responsive to habitual activity patterns (Fig. 3f).

### PPs Mediate the Relationship Between PA and Incident Health Outcomes

We evaluated associations between PA features and the risk of 14 incident health outcomes across multiple organ systems using Cox proportional hazards models adjusted for age, sex, and BMI (Supplementary Table 7). These outcomes were grouped by organ system as follows: type 2 diabetes (T2DM) representing the metabolic system; liver disease for the hepatic system; chronic obstructive pulmonary disease (COPD) for the respiratory system; ischemic heart disease (IHD) and ischemic stroke for the cardiovascular system; Alzheimer’s disease, Parkinson’s disease, amyotrophic lateral sclerosis (ALS), and vascular dementia for the neurological system; rheumatoid arthritis and inflammatory bowel disease (IBD) for the immune and inflammatory system; endometriosis and cystitis for the reproductive and urinary systems, respectively; and all-cause mortality as a general health outcome^46^.

A total of 119 significant associations were identified. The strongest protective associations were observed for Parkinson’s disease, particularly with high-intensity PA features such as MVPA (hazard ratio, HR, = 0.313, 95% CI: 0.263–0.371), MVPA bout frequency (MVPA-BF; HR = 0.332, 95% CI: 0.287–0.383), and MA (HR = 0.347, 95% CI: 0.300–0.403). Consistent associations across all PA categories were also observed for Alzheimer’s disease, vascular dementia, and COPD. All activity-related PA features exhibited protective effects on health outcomes through PPs. Notably, MVPA-BF conferred a protective effect comparable to that of average MVPA time, suggesting that frequent short bouts of activity may offer similar benefits when sustained MVPA is less feasible. Although associations with LPA were generally weaker, they remained consistently negative, supporting the potential preventive value of even low-intensity activity (Fig. 4a; Supplementary Table 8).

**Figure 4.**
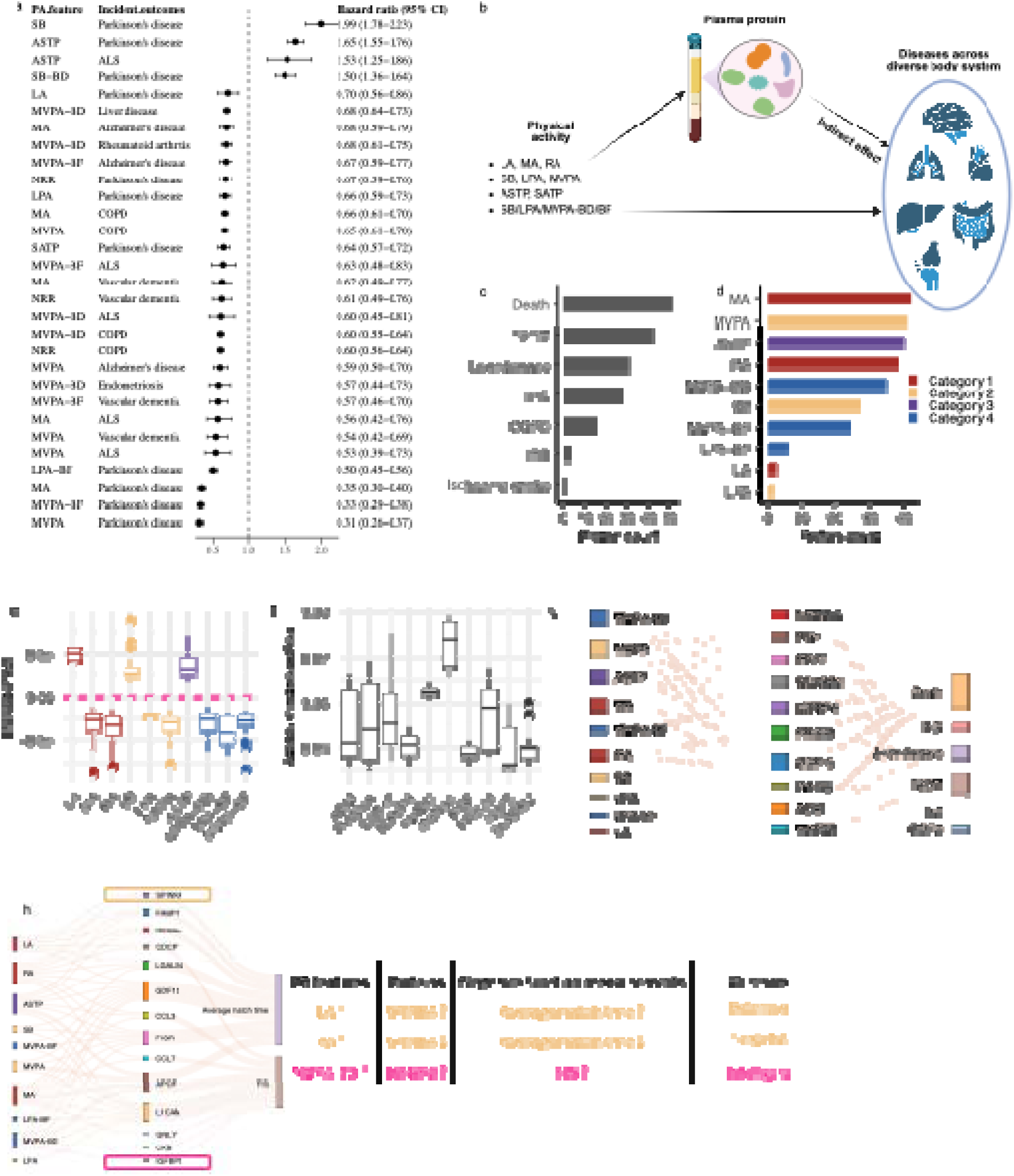
Associations between physical activity (PA) features and incident health outcomes, and mediation analysis by plasma proteins (PPs) (a) Hazard ratios and 95% confidence intervals for the associations between PA features and risk of incident health outcomes. (b) Mediation analysis framework illustrating relationships among PA features, PPs, and incident outcomes.(c) Number of PPs with significant indirect effects linked to each incident health outcome (response outcome). (d) Number of PPs with significant indirect effects linked to each PA feature (exposure). (e) Boxplot showing the indirect effects of proteins linked to each PA feature. (f) Boxplot of indirect effects of the seven PPs with the highest frequency of significant indirect effects. (g) Network diagram showing associations among PA features, the ten PPs with the most frequent or strongest significant indirect effects, and disease outcomes. (h) Network diagram showing associations among PA features, significantly associated PPs, and cognitive function measurements.

We further performed mediation analysis to evaluate whether PPs associated with PA features mediate the relationship between PA and incident health outcomes (Fig. 4b). This analysis identified 359 significant PA–PP–outcome mediation paths, involving 99 unique proteins with significant indirect effects (*p* < 1.71 × 10□□) across 10 PA features and 7 outcomes: all-cause mortality, T2DM, liver disease, IHD, COPD, IBD, multiple sclerosis, and ischemic stroke (Supplementary Tables 9–10; Fig. 4c). Among PA features, MA, ASTP, MVPA proportion, MVPA-BD, and RA were each linked to approximately 40 mediated proteins (Fig. 4d). As expected, indirect effects were consistently negative for proteins associated with higher-intensity features—MA, MVPA (proportion, BD, BF), RA, and LPA (proportion, BF)— indicating that increased activity levels, particularly MVPA, may reduce disease risk through protein-mediated mechanisms (Fig. 4e). The magnitude of indirect effects was comparable for proteins associated with MVPA-BD and MVPA-BF versus MVPA time (paired *t*-tests: *p* = 0.397, 0.425, respectively), suggesting that frequent short bouts of vigorous activity may confer benefits similar to sustained MVPA through shared molecular pathways.

To identify high impact proteins, the frequency of PPs linking PA features to incident health outcomes was evaluated. GDF15 (negatively associated with high-intensity or dynamic PA features such as MVPA-BF), a well-established biomarker ^47–49^, emerged as the most prominent mediator, involved in 24 PA–outcome paths—the highest among the 99 PPs analyzed. Six additional proteins—PRSS8, IGFBP4, SCARB2, ADM, PGF, and RBP5—each mediated at least 10 distinct paths. To identify the strongest mediators, we focused on links with absolute indirect effects (|log HR| > 0.05) and proportional mediation exceeding 10%, yielding 52 robust PA–PP–outcome paths. Beyond GDF15, ADM, PRSS8, MXRA8, FAM20A, and INHBC were notable, each mediating at least three high-impact paths (Fig. 4f–g). Notably, INHBC and MXRA8 exhibited strong indirect effects in multiple PA–T2DM paths. These findings highlight a subset of key proteins that may underlie the protective effects of physical activity and serve as candidate biomarkers of activity-induced physiological adaptation.

Our analyses confirmed several previously reported links between PA, protein expression, and health outcomes. For example, prior studies in both humans and animal models have shown that exercise elevates circulating levels of GDF15, a protein implicated in recovery from metabolic disorders such as T2DM ^50^. In line with these findings, GDF15 mediated the association between four PA features—MVPA, MVPA-BD, MA, and RA—and T2DM in our study, with an average proportional indirect effect of 13.4%. Similarly, ADM, a protein elevated in patients with COPD ^51^ and known to respond to physical activity ^52^, emerged as a significant mediator in multiple PA–COPD paths. In addition to these known molecular pathways, we identified novel protein mediators with strong indirect effects and others that acted across multiple PA–health outcomes pairs. These findings underscore both established and previously unrecognized molecular mechanisms linking PA to disease risk, offering promising targets for future biomarker discovery and intervention development.

### PPs Provide Insights into PA and Cognitive Function Associations

Multiple PA features were significantly associated with three neurological diseases, most notably Parkinson’s disease (Fig. 4a; Supplementary Table 8), prompting further investigation into links between PA-associated plasma proteins and cognitive performance. We examined associations between 613 PA-related PPs and three cognitive measures: mean time to correctly identify matches, maximum digits remembered correctly, and fluid intelligence score (FIS) (see Methods) ^53^. Fourteen unique PPs were significantly associated with at least one cognitive outcome (Bonferroni threshold: *p* < 1.71 × 10□□; Fig. 4g, Supplementary Fig. 4). Seven proteins were associated with mean time to correctly identify matches, and eight were linked to FIS, with apolipoprotein F (APOF) significantly associated with both traits. No proteins showed significant associations with maximum digits remembered correctly. These findings suggest that a subset of PA-associated proteins may influence cognitive domains relevant to neurological health.

Among the seven plasma proteins associated with mean time to correctly identify matches, six were positively associated, while APOF was negatively associated. Notably, these six proteins also exhibited negative associations with PA features related to MVPA, particularly MVPA-BF, MVPA-BD, and the proportion of MVPA time. In contrast, APOF showed negative associations with MVPA-BD, MVPA proportion, and MA, and a positive association with ASTP. These patterns suggest that MVPA may support cognitive performance by modulating levels of specific circulating proteins. Similar trends were observed for proteins linked to fluid intelligence score, with the exception of L1CAM, further reinforcing the relationship between physical activity, protein expression, and cognitive health.

### Mendelian Randomization Reveals Potential Causal Relationships Between PA-Associated PPs and Incident health outcomes

To evaluate whether the 99 plasma proteins identified as mediators may play causal roles in disease risk, we conducted Mendelian randomization (MR) analysis using genome-wide significant cis-pQTLs as instrumental variables for each protein–outcome pair ^54^. A two-sample MR framework was applied, incorporating 12 complementary methods to account for pleiotropy and heterogeneity, and supplemented with Steiger directionality testing to confirm the direction of effect from protein levels to disease risk. GWAS summary statistics for outcomes were obtained from the IEU OpenGWAS database ^55,56^.

Among the 28 plasma proteins mediating the relationship between PA features and IHD, 10 (35.7%) demonstrated potential causal effects (*p* < 1.71 × 10□□), representing the highest proportion among the seven incident outcomes assessed (Supplementary Table 11). Proteins mediating PA pathways to all-cause mortality, liver disease, and T2DM also frequently showed evidence of causality, with 10 (19.2%), 5 (16.1%), and 12 (28.6%) proteins, respectively. In contrast, among the 10 proteins mediating PA–COPD associations, only one, *CHCHD10*, exhibited a significant causal effect. These findings suggest that differential molecular mechanisms underlie the relationship between physical activity and health outcomes, with stronger genetic evidence supporting causality in cardiovascular and metabolic pathways.

Our analysis confirmed several previously reported causal protein–health outcomes relationships (Fig. 5a,b). Furin, a proprotein convertase involved in the activation of multiple precursor proteins, including pro-inflammatory cytokines and growth factors, was causally linked to IHD. Its role in vascular remodeling and plaque instability has been well documented in the context of atherosclerosis ^57^. WFDC2, also known as HE4, is a secreted glycoprotein implicated in innate immunity and protease inhibition. WFDC2 is overexpressed in multiple malignancies, including HCC, and elevated serum levels have been proposed as a biomarker for early detection and prognosis^58^. Its association with health outcomes progression suggests potential for therapeutic targeting. INHBC, a member of the TGF-β superfamily, was identified as a significant mediator in PA–T2DM paths and has been implicated in T2DM through both proteomic and genetic studies. A large-scale proteomic screen involving 4,775 proteins identified INHBC as associated with incident T2DM ^59^, with this relationship further supported by Mendelian randomization analysis^60^.

**Figure 5.**
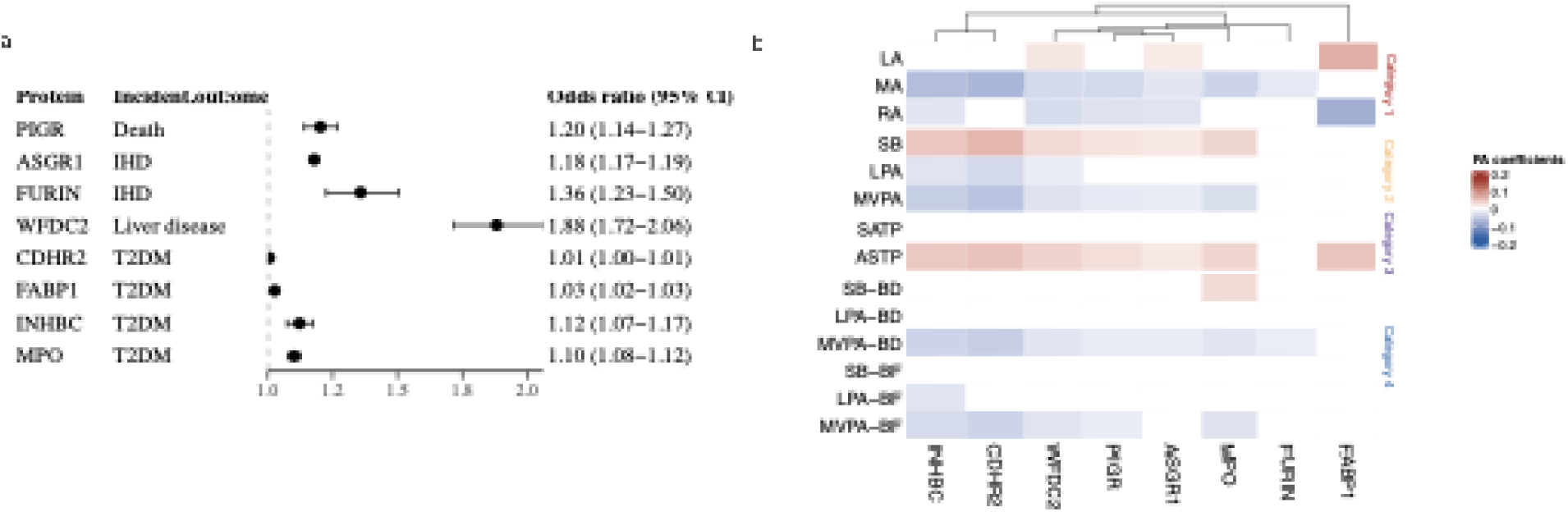
Genetic analysis results. (a) Mendelian randomization results (odds ratio) of plasma proteins identified as mediators of PA–disease associations that also demonstrated potential causal effects on disease outcomes **(b)** Heatmap showing the linear model coefficients between PA features and PPs with evidence of causal effects on disease outcomes.

Bayesian colocalization analysis was performed to evaluate whether the same genetic variant influences both plasma protein levels and disease risk^61^. Among the proteins mediating associations between PA features and type 2 diabetes, 25 of 42 (59.5%) showed moderate-to-high colocalization (posterior probability for shared causal variant, PP.H4 = 0.80–0.99; Supplementary Table 12). Notably, seven proteins—FCAMR, GSTA1, ACY1, ADH4, HAO1, CA5A, and SERPIND1—exhibited near-complete colocalization (PP.H4 ≈ 1). A similar pattern was observed for ischemic heart disease, where 15 of 28 (53.6%) mediating proteins showed moderate-to-high colocalization. In contrast, only 4 of 31 (12.9%) proteins mediating associations with liver disease showed evidence of colocalization, and no colocalization was detected for proteins mediating pathways to all-cause mortality or COPD. These findings suggest a stronger genetic basis for the protein-mediated effects of PA on metabolic and cardiovascular outcomes compared to other disease domains.

We identified 8, 6, and 1 PPs that exhibited both causal effects and shared genetic variants with T2DM, IHD, and liver disease, respectively. While colocalization strengthens causal inference, the absence of colocalization does not preclude causality, as complex genomic loci may contain multiple independent signals^62^. Nonetheless, these findings suggest that a substantial subset of PA-associated PPs possess strong genetic evidence for causal involvement in disease risk. This highlights their potential as therapeutic targets and warrants further functional investigation.

## Discussion

In summary, among more than 9,000 participants with data from both the UK Biobank accelerometry substudy and the Olink Explore 3072 proteomics platform, we quantified associations between 14 accelerometer-derived PA features—spanning four categories: average intensity, proportional activity status, TP, and bout characteristics—and PP expression. Our findings reveal that PP expression levels are linked to multiple behavioral dimensions of PA. Some proteins, such as ITGAV and MYOM3, were associated with all PA categories, while others demonstrated specificity for particular aspects, such as the duration of MVPA. The protein-coding genes linked to distinct PA patterns are significantly enriched in pathways governing immune modulation, cellular growth, and cardiac remodeling. Foremost among these is the PI3K-Akt cascade, a highly conserved intracellular signalling hub that orchestrates cell survival, metabolism, and proliferation. Notably, PI3K-Akt enrichment emerges across the full spectrum of PA behaviours, from sedentary phenotypes (e.g., ASTP, SB) to more vigorous patterns (e.g., MVPA-BF, MA, MVPA). This ubiquity mirrors the pathway’s well-established involvement in both physiological adaptation and pathological remodelling of multiple tissues. Finally, the plasma proteomic signatures likely capture proteins released from several organs; indeed, many PA-associated proteins (such as MMP8 and MMP9 enriched in lung) show tissue-selective expression patterns in the enhanced GTEx consortium dataset ^36–38^, underscoring their systemic origin and potential as interorgan biomarkers of activity-dependent signalling.

Risk of incident health outcomes was significantly associated with all four categories of PA features, with more vigorous activity-related features consistently demonstrating protective effects. Notably, one exception to the overall trend was ALS. While previous studies have reported mixed findings—some suggesting PA as a potential risk factor^63^ and others reporting no clear association^64^—we observed a protective association between PA and ALS risk in our analysis. This discrepancy may be partly explained by the context of our study: participants engaged in physical activity within a free-living environment, rather than in response to imposed or structured exercise protocols. In this setting, higher levels of activity may serve as a proxy for an overall healthier lifestyle, which could confound the observed association.

Mediation analyses revealed that several PPs, including GDF15, PRSS8, and IGFBP4, partially mediated the associations between PA features and health outcomes, with some mediators spanning multiple PA categories. Notably, bout-related features such as MVPA-BF and MVPA-BD exhibited indirect effects comparable in magnitude to those of proportional MVPA time, suggesting that frequent short bouts of activity may offer similar health benefits to sustained exercise. Mendelian randomization analyses further supported potential causal roles for several of these mediator proteins in influencing disease risk.

While previous studies have examined associations between PA and PP expression, they have often been constrained by limited sample sizes, narrow proteomic coverage, restricted PA feature sets, or a limited range of health outcomes^11–13^. In contrast, our study systematically integrates a large-scale proteomics platform with 14 PA features spanning four behavioral categories, offering a detailed characterization of habitual activity patterns. We complemented this with comprehensive protein annotation analyses to elucidate the biological functions of PA-associated PPs. Furthermore, by incorporating two-sample Mendelian randomization and colocalization analyses, we assessed the potential genetic links between PPs and disease outcomes, providing insights into their therapeutic relevance.

Several limitations warrant consideration. First, our analysis was cross-sectional, and although we assumed that PA features are more likely to influence PP concentrations, reverse causality or bidirectional relationships cannot be excluded. Second, accelerometry data were collected during a single 7-day period, limiting our ability to assess temporal variability in PA behavior. Longitudinal accelerometry would enable the study of dynamic changes in activity and their molecular correlates over time. Lastly, our findings may have limited generalizability, as the majority of participants were white individuals of European ancestry.

Our findings suggest that shorter, more frequent bouts of MVPA are associated with health benefits comparable to those of overall MVPA time. MVPA-BF also exhibited indirect effects of similar magnitude in protein-mediated paths, underscoring the potential health benefits of accumulating activity through multiple short bouts, especially when sustained exercise is not feasible. Building on these insights, future work could integrate all categories of PA features into unified machine learning models to evaluate their joint and independent contributions to disease risk. Such models could quantify the specific impact of bout frequency while accounting for total activity duration across intensity levels. Ultimately, this approach may inform the development of personalized, data-driven recommendations to promote healthier lifestyles and reduce the burden of chronic disease.

## Methods

### UKB Participants

The UKB is a population-based prospective cohort comprising approximately 500,000 participants aged 40–69 years at recruitment (2006–2010), with follow-up extending up to 18 years ^65^. The dataset includes de-identified genetic, lifestyle, and health-related information, as well as biological samples. While the cohort is not demographically representative of the global population, its large sample size and extensive phenotypic diversity enable robust investigation of a wide range of exposure–outcome associations ^66^.

### UKB Accelerometry Substudy

A subset of UK Biobank participants was invited to participate in the accelerometry substudy, in which individuals were asked to wear a triaxial wrist-worn accelerometer continuously for up to 7 days ^16,67^. This lightweight device records tri-axial acceleration signals to estimate movement intensity. Of the 236,279 individuals invited, 132,661 either declined or did not respond, resulting in approximately 100,000 participants with at least three days of valid accelerometer data^68^.

The UKB provides both raw accelerometry data and derived PA features, including average acceleration measured in milligravitational units (milli-g), recorded in 5-second epochs. To reduce computational burden and minimize noise, we aggregated the 5-second data into 1-minute intervals. From these aggregated data, we derived 14 PA features across four categories: average intensity, proportional activity status, TP, and bout characteristics. Average log acceleration was computed for the LA 6-hour period (00:00–06:00), typically corresponding to sleep, and the MA 18-hour period (06:00–00:00). RA was then calculated as (MA – LA) / (MA + LA), reflecting the contrast between rest and activity and enabling assessment of their joint influence on plasma protein expression.

Each minute of accelerometry data was classified as SB, LPA, or MVPA minute based on mean acceleration thresholds: SB was defined as <30 milli-g, MVPA as >100 milli-g, and LPA as 30–100 milli-g ^17^. To account for differences in total wear time across participants, we calculated the proportion of time spent in each activity category.

The ASTP was defined as the probability that a given minute was classified as SB following a minute of LPA or MVPA^17^. Conversely, the SATP quantified the likelihood of transitioning from SB to either LPA or MVPA. Bout duration was defined as the mean length of uninterrupted SB, LPA, or MVPA episodes, while bout frequency captured the average number of such episodes per day ^18^.

### UKB Plasma Protein Project (UKB-PPP)

Proteomic profiling was performed on blood samples from 54,219 UK Biobank participants using the antibody-based Olink Explore 3072 proximity extension assay (PEA), which quantified 2,923 unique PPs. The cohort comprised 46,595 (85.9%) randomly selected individuals, 6,376 (11.8%) selected by a consortium, and 1,268 (2.3%) participants from a COVID-19 repeat imaging study. To minimize potential confounding effects of COVID-19, we restricted analyses to samples from the first two groups, collected between March 2006 and September 2010^10,69^.

A subset of participants was further recruited between 2015 and 2019, and again in 2021, for repeated plasma protein measurements. However, due to the limited sample size (n = 1,173) and the small overlap with the PA cohort (n = 506), these repeated measurements were excluded from downstream analyses.

### Covariates

The UKB collected comprehensive sociodemographic, lifestyle, and medical history data via touchscreen and in-person questionnaires. In our analysis, we initially considered four commonly used confounders: age, sex, race, and BMI ^10^. However, due to the extremely limited number of non-white participants in the final cohort (n = 2), race was excluded as a covariate in subsequent analyses.

### Preprocessing

Following the UKB accelerometer data preprocessing protocol, we excluded participants with poor-quality data, insufficient wear time for averaging activity profiles, or uncalibrated devices ^68^. This yielded 96,504 participants with “good” accelerometer data, as defined by UKB quality control criteria. We further excluded individuals with missing values for PA features, resulting in a final sample of 87,716 participants with complete data. All continuous PA features, as well as BMI and age, were standardized to z-scores prior to analysis.

Good-quality PA data were available for 9,210 participants (17.0%) within the proteomics cohort. We assessed associations between PA features and plasma protein concentrations by fitting separate linear models for each PA feature–protein pair. Participants with missing protein concentration values were excluded on a per-model basis. On average, each model included 8,152 participants (SD = 759). One protein, GLIPR1, had data available for only 30 individuals—a sample size markedly below the cohort average—and was therefore excluded from further analysis.

### PA and PP association analysis

Scaled PA features were used as predictors, and inverse-rank normalized PP expression levels were used as outcomes in linear models adjusted for age, sex, and BMI. All statistical analyses were performed using R (version 4.3.3). Statistical significance was defined as *p* ≤ 1.71 × 10□□, applying a Bonferroni correction for 2,922 protein tests.

### Biological function analysis of PA-associated PPs

To investigate the potential biological functions of PA-associated PPs, we conducted pathway and tissue enrichment analyses. For each set of proteins significantly associated with a given PA feature, pathway enrichment analysis was performed using the GO:BP and KEGG databases ^29,30,32^. FDR correction was applied to identify significantly enriched biological processes and pathways. For GO:BP, we additionally identified key driver terms to highlight core functional themes within the enriched gene sets ^70^.

Tissue enrichment analysis of PA-associated PPs was performed using tissue-enriched gene expression data from the GTEx database, which includes approximately 12,000 genes across 32 normal human tissues ^38^. These tissues were grouped into 15 biologically defined systems, including the brain, heart, skin, intestine, endocrine system, tibial nerve, artery, lung, pancreas, liver, breast, spleen, skeletal muscle, and sex-specific tissues (gynecotropic and androtropic), as detailed in Supplementary Table 6. For each system, the final set of tissue-enriched proteins was defined as the union of enriched proteins from all constituent tissues. PA-associated PPs were then intersected with each tissue-specific set to evaluate enrichment across systems for all 14 PA features spanning four categories. Fisher’s exact test was applied to determine whether the proportion of tissue-enriched PPs associated with a given PA feature was significantly elevated within each tissue system. This analysis was performed independently for each PA feature.

### Mediation analysis

We examined the potential mediating role of PPs in linking PA features to incident health outcomes using mediation analysis ^71^. Associations between each PA feature and the risk of 14 incident health outcomes were first assessed using Cox proportional hazards models, adjusted for sex, age, and BMI. The outcomes included all-cause mortality, T2DM, liver disease, COPD, IHD, Alzheimer’s disease, ALS, Parkinson’s disease, ischemic stroke, vascular dementia, rheumatoid arthritis, IBD, endometriosis, and cystitis. Owing to limited case numbers following PA data collection, some outcomes comprised multiple disease subtypes. Disease classification followed the UK Biobank disease categorization scheme, informed by clinical expertise. Detailed categorization criteria and corresponding ICD-10 codes are provided in Supplementary Table 7.

Follow-up time and data on incident health outcomes were obtained from the UK Biobank. For each outcome, follow-up began at the baseline of the accelerometry substudy (2013–2015) and continued until the earliest of outcome diagnosis or the study end date, September 1, 2023. Incident outcomes that showed statistically significant associations with PA features—adjusted for multiple comparisons using the Benjamini Hochberg procedure—were selected for downstream mediation analysis.

Within the mediation framework, each PA feature was treated as the exposure variable, and PPs significantly associated with that feature were considered potential mediators. Incident health outcomes, along with their timing, served as the outcome variables. All models were adjusted for age, sex, and BMI. Indirect effects were estimated using the conventional bootstrap method with 1,000 resamples to evaluate statistical significance ^72^. PPs with significant indirect effects (*p* < 1.71 × 10□□) were classified as mediating proteins linking PA features to incident health outcomes.

### Association with Cognitive Functions

Cognitive tests assessing processing speed, reaction time, reasoning, and memory were administered on the same day as participants’ initial assessment visit. Linear models were used to evaluate associations between PA-associated PPs and three cognitive performance measures: mean time to correctly identify matches, maximum digits remembered correctly, and FIS. Models were adjusted for age, sex, and BMI. Associations with *p* ≤ 1.71 × 10□□ were considered statistically significant, applying a Bonferroni correction for multiple testing.

### Mendelian randomization analysis

We applied conventional two-sample MR methods to investigate causal relationships between identified mediating PPs and incident health outcomes, using genetic variants as instrumental variables. Instruments were selected as genome-wide significant (*p* < 5 × 10□□) cis-pQTLs located within ±1 Mb of the gene, based on data from the UKB-PPP study ^10^. Variants were clumped for linkage disequilibrium (LD) using PLINK (version 1.9) with an *r²* threshold of <0.01 and the 1000 Genomes European reference panel. To reduce weak instrument bias, variants with an F-statistic <10 were excluded.

GWAS summary statistics for each incident health outcome were obtained from publicly available resources within the IEU GWAS databases, incorporating data from large-scale cohorts including the NHGRI-EBI GWAS Catalog ^73^, MRC IEU OpenGWAS ^55^, and FinnGen ^74^, restricted to individuals of European ancestry. Two-sample MR analyses were conducted using the *TwoSampleMR* package in R ^56,75^.

### Colocalization Analysis

A Bayesian colocalization approach was used to detect shared causal variants between protein cis-pQTLs (UKB-PPP ^10^) and GWAS hits for selected incident health outcomes^61^. The GWAS data were retrieved from the OpenGWAS database, specifically focusing on the European cohort

Significant loci for PPs were defined using pQTL signals with a significance threshold of 5.0 × 10□□. These loci were LD-clumped with an r² < 0.1 within a ±1 Mb window of the lead variant. Loci with at least 50 overlapping SNPs between pQTL and GWAS summary statistics and at least one genome-wide significant SNP (p < 5.0 × 10□□) were tested using the R coloc package with default priors (p1 = 1 × 10□□, p2 = 1 × 10□□, p12 = 1 × 10□□). A posterior probability of H4 greater than 0.8 was considered indicative of a colocalized signal, suggesting that the PP and the incident health outcome share a common associated or causal variant.

## Data availability

All UK Biobank data (accelerometer, demographics, proteomic) are accessible via application to the UK Biobank. PQTL summary statistics from the UKB-PPP can be accessed at https://metabolomips.org/ukbbpgwas.

## Code availability

All analyses and visualization were implemented in R (v4.4.0). Scripts are available upon reasonable request, including those used to replicate the analyses and figures presented in this manuscript.

## Acknowledgements

This study utilized data from the UK Biobank resource under application numbers 197947. We sincerely thank the participants and staff of the UK Biobank for the invaluable contributions. M.W., H.Q., and Y.Y were supported by M.W.’s startup fund from the Institute of Heart and Brain Health. This work was supported by grants the NIH R35 HL155318 to A.R. and the American Heart Association (AHA) grants 23MERIT1038415 to A.R. and SFRN (URLs: https://doi.org/10.58275/AHA.24SFRNPCN1284382.pc.gr.194135 and https://doi.org/10.58275/AHA.24SFRNCCN1276092.pc.gr.194131) to A.R. and M.W.

## Author contributions

M.W. conceived the study, supervised the research, and revised the manuscript. A.R. revised the manuscript and supervised the research. H.Q. performed the analyses and wrote the manuscript. C.W. supported pathway and biological process analyses and revised the manuscript. Y.Y. preprocessed the data. B.L. revised the manuscript and figure preparation.

---

**Supplementary Figure 1.**
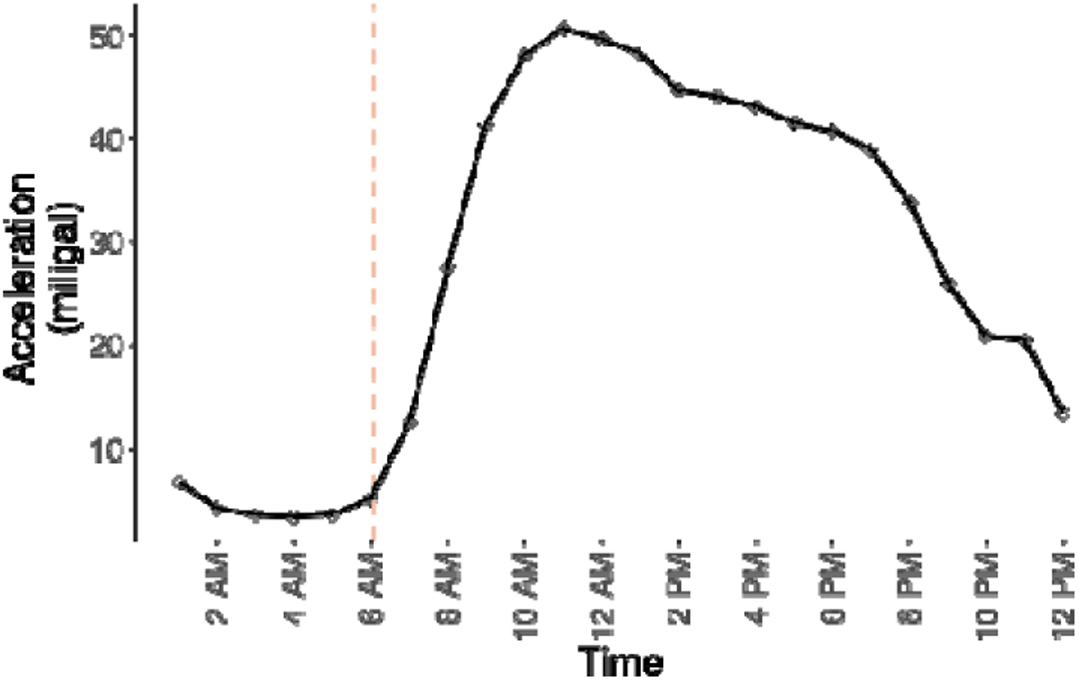
Average acceleration displayed in one-hour windows across 24 hours, showing participants exhibit lower activity levels from 12:00 AM to 6:00 AM.

**Supplementary Figure 2.**
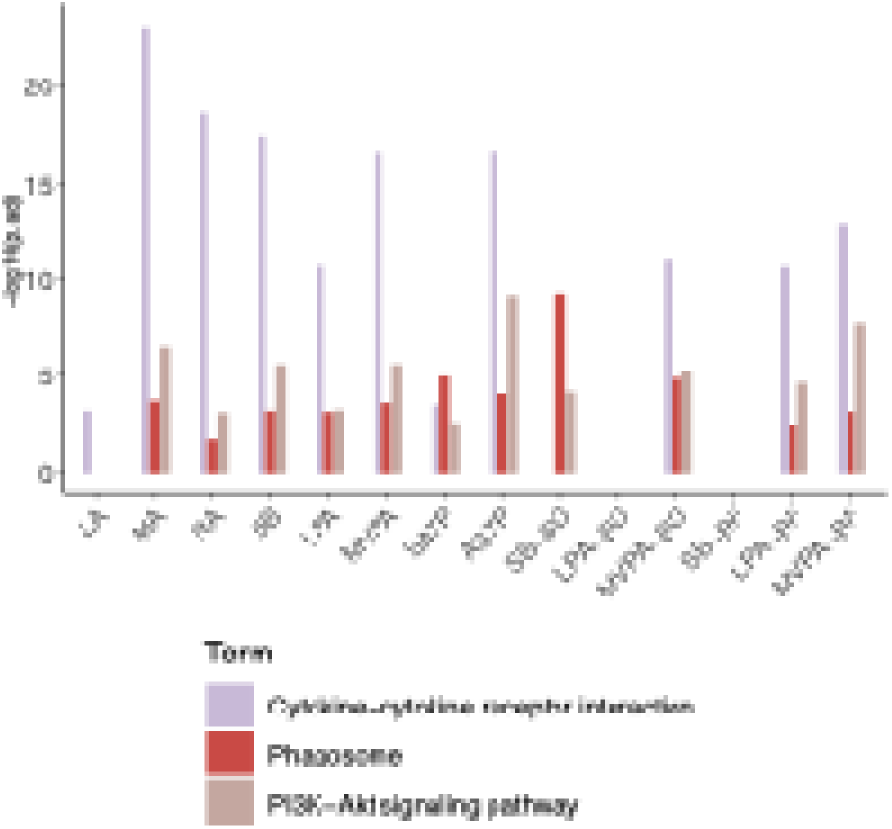
Pathway enrichment analysis results indicted three biological pathways (cytokine-cytokine receptor interaction, phagosome, and PI3K-Akt signaling pathway) commonly enriched by PPs associated with distinct PA features.

**Supplementary Figure 3.**
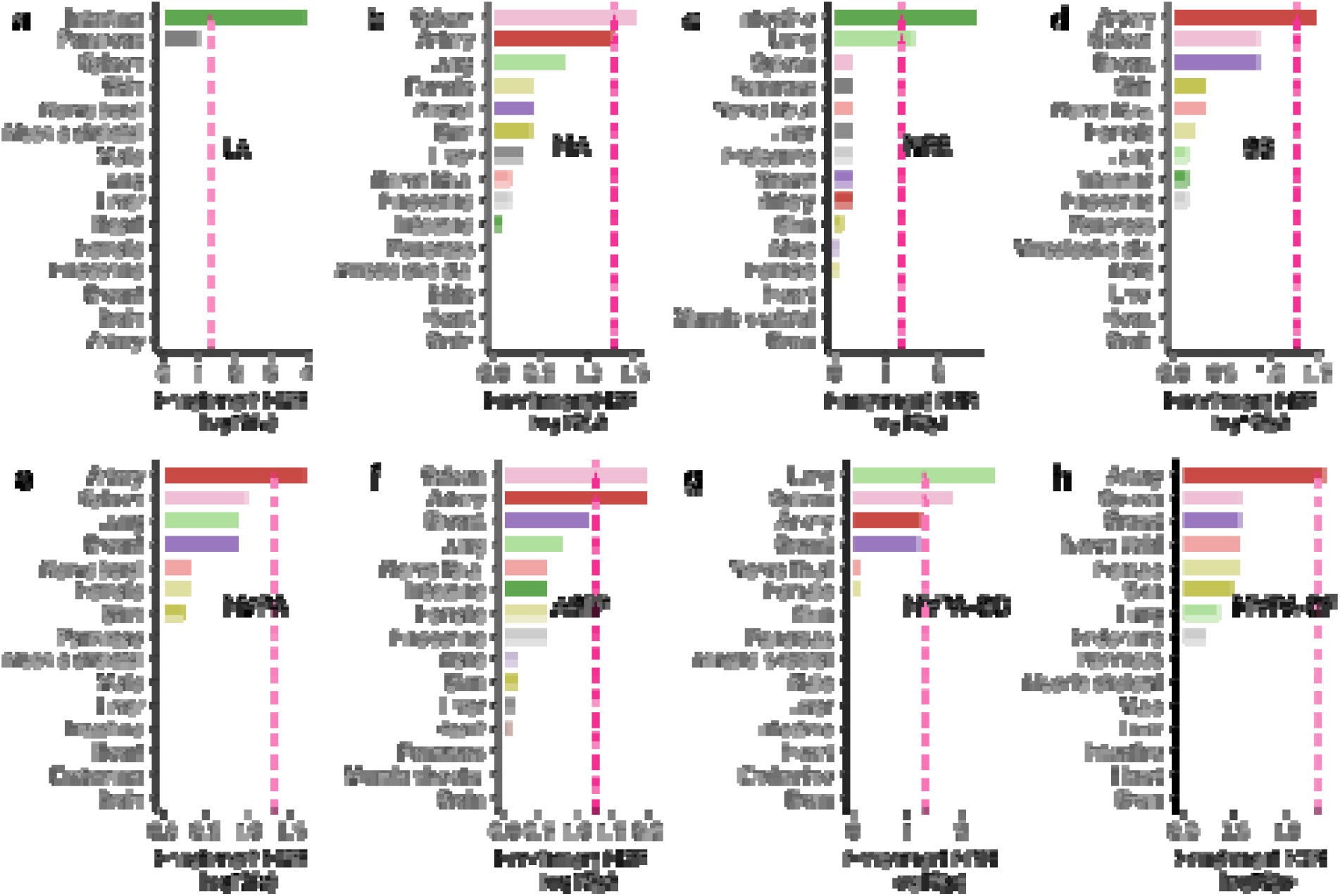
Fisher’s exact test results showing the enrichment of plasma proteins associated with each PA feature across distinct organ systems. Only protein sets with significant tissue-specific enrichment (Benjamini–Hochberg adjusted *p* < 0.05) are displayed.

**Supplementary Figure 4.**
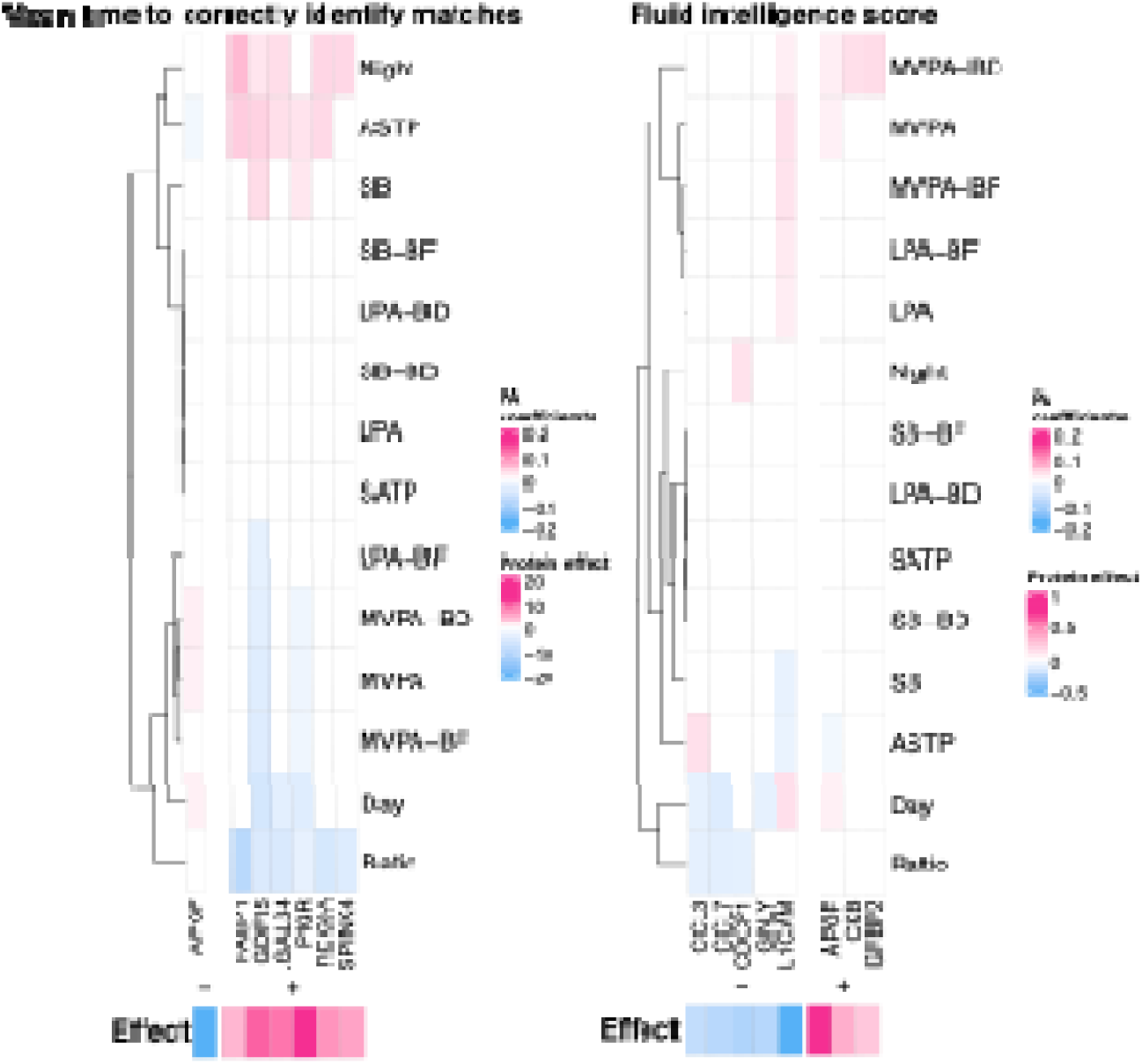
Heatmaps of coefficients for plasma proteins (PPs) associated with physical activity (PA) features and cognitive performance measures: (a) mean time to correctly identify matches and (b) fluid intelligence score. The main heatmap displays associations between PA features and PPs, while the bottom bar indicates the direction and strength of associations between the corresponding PPs and cognitive outcomes. Red indicates a positive association; blue indicates a negative association.

